# Extended High-Frequency Audiometry using the Wireless Automated Hearing Test System Compared to Manual Audiometry in Children and Adolescents

**DOI:** 10.1101/2023.05.22.23290339

**Authors:** Chelsea M. Blankenship, Lindsey M. Hickson, Tera Quigley, Erik Larsen, Li Lin, Lisa L. Hunter

## Abstract

**Objectives:** Reliable wireless automated audiometry that includes extended high frequencies (EHF) outside a sound booth would increase access to monitoring programs for individuals at risk for hearing loss, particularly those at risk for ototoxicity. The purpose of the study was to compare thresholds obtained with 1) standard manual audiometry to automated thresholds measured with the Wireless Automated Hearing Test System (WAHTS) inside a sound booth, and 2) automated audiometry in the sound booth to automated audiometry outside the sound booth in an office environment.

**Design:** Cross-sectional, repeated measures study. Twenty-eight typically developing children and adolescents (mean = 14.6 yrs; range = 10 to 18 yrs). Audiometric thresholds were measured from 0.25 to 16 kHz with manual audiometry in the sound booth, automated audiometry in the sound booth, and automated audiometry in a typical office environment in counterbalanced order. Ambient noise levels were measured inside the sound booth and the office environment were compared to thresholds at each test frequency.

**Results:** Automated thresholds were overall about 5 dB better compared to manual thresholds, with greater differences in the extended high frequency range (EHF;10-16 kHz). The majority of automated thresholds measured in a quiet office were within ± 10 dB of automated thresholds measured in a sound booth (84%), while only 56% of automated thresholds in the sound booth were within ± 10 dB of manual thresholds. No relationship was found between automated thresholds measured in the office environment and the average or maximum ambient noise level.

**Conclusions:** These results indicate that self-administered, automated audiometry results in slightly better thresholds overall than manually administered audiometry in children, consistent with previous studies in adults. Ambient noise levels in a typical office environment did not have an adverse effect on audiometric thresholds measured using noise attenuation headphones. Thresholds measured using an automated tablet with noise attenuating headphones could improve access to hearing assessment for children with a variety of risk factors. Additional studies of extended high frequency automated audiometry in a wider age range are needed to establish normative thresholds.

## INTRODUCTION

The recent World Report on Hearing estimates that 34 million children around the world are living with a hearing loss that is affecting their health and quality of life (<u>World Health Organization., 2021</u>). Hearing loss in children, even slight to mild in degree, is associated with a variety of developmental differences (<u>Moore et al., 2020</u>). Delayed speech and language skills, social problems, and struggle in academic settings are common adverse consequences of pediatric hearing loss (<u>Joint Committee on Infant Hearing., 2019</u>). The primary method of combating these effects is through increased access to screening and diagnostic hearing healthcare.

The Joint Committee on Infant Hearing guidelines outline twelve categories of risk factors for infants that pass the newborn hearing screen that should be monitored for progressive or late onset hearing loss as well as recommended diagnostic follow-up (<u>Joint Committee on Infant Hearing., 2019</u>) Postnatal risk factors included in the Joint Committee on Infant Hearing guidelines are “culture-positive infections associated with sensorineural hearing loss including confirmed bacterial and viral meningitis or encephalitis, events associated with hearing loss such as significant head trauma especially basal skull/temporal bone fractures and chemotherapy, and caregiver concern regarding hearing, speech, language, development delay and or developmental regression” (<u>Joint Committee on Infant Hearing., 2019</u>). These late-onset hearing losses are underdiagnosed, but are typically detected by pediatrician or a school hearing screening.

Another group of individuals at high risk for hearing loss are those receiving aminoglycoside antibiotics that are potentially toxic to the inner ear. Ototoxicity is the irreversible degradation of auditory function resulting from the physiologic effects of antibiotics on the sensory hair cells within the cochlea (<u>American Academy of Audiology, 2009</u>). Most commonly, damage is initially evident in the high frequency regions and then progresses to the lower frequencies, ultimately affecting the person’s ability to understand speech (<u>Al-Malky et al., 2011</u>; <u>Blankenship et al., 2021</u>; <u>Fausti et al., 1992</u>; <u>Garinis et al., 2021</u>; <u>Garinis et al., 2018</u>). Hearing monitoring programs traditionally only include frequencies up to 8 kHz, even though the measurement of extended high frequency thresholds (EHF; 8-16 kHz) has long been recommended for ototoxicity monitoring (<u>American Academy of Audiology, 2009</u>; <u>American Speech-Language-Hearing Association, 1995</u>; <u>Fausti et al., 1992</u>). EHF audiometry is more sensitive to outer hair cell damage that occurs as a result of ototoxicity, however access is limited, especially for serial monitoring.

The ability to automatically test EHF hearing outside of the sound booth is a relatively new development. Recent published data in children and adults is available using portable automated testing for frequencies from 0.25 to 8 kHz (<u>Bastianelli et al., 2019</u>; <u>Magro et al., 2020</u>; <u>Meinke et al., 2017</u>; <u>Serpanos et al., 2022</u>; <u>Thompson et al., 2015</u>; <u>Whitton et al., 2016</u>).Additionally, <u>Clavier et al. (2022)</u> demonstrated the validity of standard and EHF thresholds obtained using the Wireless Automated Hearing Test System (WAHTS) in adults. However the validity of automated thresholds obtained with the WAHTS have not yet been examined in children, specifically within the EHF region. EHF audiometry employs circumaural headphones that are calibrated for the regions above 8 kHz. Existing ototoxicity guidelines state that it is best practice to measure EHF thresholds in a sound booth rather than to use a portable audiometer at the bedside or other patient settings (<u>American Academy of Audiology, 2009</u>; <u>Fausti et al., 1993</u>). For EHF thresholds to be reliably measured, a clinician must have the available equipment and the patient must be healthy enough to visit a sound booth. However, portable audiometers that are automatic, valid outside a sound booth, and can test frequencies higher than 8 kHz have the potential to make ototoxicity monitoring more accessible for children and adults who are confined to hospital rooms. Mobile technology options have been developed and, while not yet standard of care, could be considered as an alternative to traditional audiometric booth testing (<u>Brungart et al., 2018</u>). Thus, the need for research in wireless and booth-less automated audiometry is clear, particularly in the pediatric population (<u>Cheng et al., 2009</u>).

Ambient noise measurements need to be recorded and taken into consideration when hearing tests are performed outside the booth to determine the accuracy of the test. If noise levels become too loud in the testing environment, audiometric thresholds can be elevated due to acoustic masking or distraction from the test tone. Maximum permissible ambient noise levels for the inside of a sound booth using supra-aural earphones should be no louder than 37 dB SPL in the frequency range of 0.25-8 kHz (ANSI S3.1-1999). The Occupational Safety and Health Administration recommends that when testing hearing in an open room, noise levels should be monitored throughout the entire session (<u>Meinke et al., 2017</u>).

This study had two specific aims designed to compare audiometry results in children, teens, and young adults. The first aim was to compare automated thresholds measured with the WAHTS inside a sound booth to manual thresholds obtained with a clinical audiometer. The second aim was to compare automated thresholds obtained within the sound booth with automated thresholds obtained in an office setting (non-sound treated room). The hypothesis was that an automated system with passive ambient noise reduction headphones and automated tracking audiometry would provide reliable thresholds independent of the test environment and that automated thresholds would be comparable to standard manual audiometry performed in a sound booth.

## METHODS

### Participants

This research was part of a larger longitudinal study examining the onset, progression and factors associated with hearing loss in children and adults with cystic fibrosis, being treated with aminoglycoside antibiotics. For the present report, typically developing children, teens and young adults were recruited from website advertisements. Children were not excluded due to hearing concerns, hearing loss, or middle ear dysfunction in order to include a wider range of hearing levels. Additional eligibility criteria included the ability to complete a behavioral hearing assessment. The study was approved by the Institutional Review Board at the hospital. Informed parental consent and child assent (for ages 11 to 17 years) or participant consent (ages 18 and older) was obtained prior to participation in the study. All participants were paid for participation.

### Procedures

Testing typically lasted 2 hours and was completed over the course of one or two visits. A participant and parental report hearing and balance history and symptom questionnaire (<u>Blankenship et al., 2021</u>) was administered to determine if the participant reported any hearing difficulties, tinnitus, balance disturbance, history of otitis media, PE tubes, or previous hearing exams. Otoscopy was performed to ensure the ear canal was patent and if necessary, cerumen was removed. Tympanometry at 226 Hz was performed using the Interacoustics Titan PC-controlled immittance system (Middlefart, Denmark). These tests were completed to detect otologic problems, but were not analyzed specifically for this report.

### Manual Audiometry

Manual audiometric thresholds were measured using the Interacoustics Equinox 2.0 with Sennheiser HDA 300 (Wennebostel, Germany) circumaural headphones. Thresholds were measured at octave test frequencies in the standard frequency range (0.25 to 8 kHz) and in the EHF range (10, 12.5, 14 and 16 kHz) using the modified Hughson-Westlake method (down 10 dB, up 5 dB) of pure-tone presentation (<u>Carhart & Jerger, 1959</u>). Bone conduction thresholds were measured with the manual test method if air conduction thresholds were ≥ 20 dB HL using a Radio-ear B-71 bone oscillator (Radioear Corp, New Eagle, PA) at 0.25, 0.5, 1, 2, and 4 kHz with appropriate narrowband masking in the contralateral ear, in order to classify type of hearing loss.

### Automated Audiometry

Automated audiometry was performed using the WAHTS system and TabSINT software provided by Creare LLC (Hanover, NH) (<u>Clavier et al., 2022</u>; <u>Shapiro et al., 2020</u>). The system consists of a tablet (Samsung SM-T377A Galaxy Tab E) and wireless headset that are connected via Bluetooth. The headset includes a wireless audiometer circuit including a digital signal processor, speaker and microphone, and noise control components such as large attenuating earcups, faceplate and protective fabric, and ear seal. The headset provides high levels of passive attenuation of approximately 30 to 40 dB, which is equivalent or better than attenuation within a sound booth. For further detail on the WAHTS, TabSINT software, calibration and RETSPL values, the reader is referred to (<u>Brungart et al., 2018</u>; <u>Clavier et al., 2022</u>; <u>Meinke et al., 2017</u>).

Prior to testing, the participant was given the tablet and prompted to read the test instructions displayed on the screen. Verbal re-instruction was delivered if the participant seemed unsure or needed clarification, but this was rarely necessary. Stimuli were presented with a Békésy-like tracking algorithm (fixed frequency) and thresholds were measured in ascending order from 0.25 to 16 kHz, the same order as manual audiometry. While taking the test, the participant was able to view the test frequency, the ear that stimuli were being presented to, and a large red response button. An example of the screen design can be seen as a figure in Supplemental Digital Content 1, with participant instructions shown on the left and the test screen on the right.

### Test Environments and Order

Automated audiometry was completed in two different test environments, a sound booth and a typical office space, while manual audiometry was only completed in the sound booth. Two single room, double-walled controlled acoustical environments (Industrial Acoustics Company, Inc. Model 120A) were used for manual and automated audiometry. The office space was selected as a relatively quiet but not acoustically-controlled environment in order to provide a realistic test of a non-sound booth test environment. The office space was carpeted, had acoustic ceiling tiles and a large conference table in the middle of several cubicles. During testing, the room was occupied by employees as well as those passing through the room to enter connected office spaces. The employees were aware that testing was occurring but were not instructed to act differently when testing was being conducted. For all tests, the ear (left vs. right) and audiometry method (manual vs. automated) order was counterbalanced so that half of the participants completed testing in their right ear first and half the participants completed manual audiometry first.

### Ambient Noise Measurements

A Larson-Davis system 824 sound level meter (Depew, New York) with a Brüel & Kjær half-inch free field microphone (type 4189, Nærum, Denmark) was used to measure ambient noise levels for each test location. Measurements included the long-term average (Leq), minimum and maximum levels (dBA SPL). For the office environment, sound level measurements were taken throughout each testing session due to the possibility of fluctuating acoustic levels. Sound level measurements were only recorded once within the sound booth since it is a controlled acoustical environment. The sound booth recording continued until the Leq stabilized.

### Statistical Analysis

Descriptive statistics were used to summarize sample demographics, results of a hearing and balance history and symptom questionnaire, and audiometric thresholds across test frequency (0.25 to 16 kHz), method and location (hereinafter referred to as “method”; manual sound booth, automated sound booth, automated office). Separate linear mixed models were conducted to evaluate threshold differences across test frequency for manual vs. automated audiometry in the sound booth and automated audiometry in the sound booth compared to the office environment. The interaction between method and frequency was explored. Tukey-Kramer adjustment was applied for multiple comparisons on significant factors. Intra-class correlation coefficients were computed to evaluate the threshold agreement and degree of correlation among methods for each frequency. Spearman rank correlations were used to assess the relationship between automated thresholds measured in the office and ambient noise levels (Leq and maximum sound pressure level). All data were collected and managed using REDCap, a secure web-based research database platform (<u>Harris et al., 2019</u>; <u>Harris et al., 2009</u>), exported and analyzed using JASP version 0.14.1.0 (open source statistical analysis program). Two-sided significance level was set at *p* < 0.05.

## RESULTS

After exclusion of one individual with a foreign body in the ear canal, a total of twenty-eight children and adolescents participated in the study. The mean age at test was 14.6 years (range = 10.2 to 18.8 yrs); 50% were males, and 59% were Caucasian. The hearing and balance history and symptom questionnaire showed 7% (n = 2 participants) reported hearing difficulties, 4% (n = 1) had tinnitus, 11% (n = 2) experienced dizziness, 18% (n = 5) had a history of otitis media with PE tubes, and 86% (n = 24) had a previous hearing exam. Audiometry analysis included one ear per participant (n = 28 ears) with automated audiometry completed in both locations (sound booth and office environment) as well as manual audiometry in the sound booth.

### Manual vs. Automated Audiometry in the Sound Booth

Manual and automated thresholds in the sound booth are shown in Figure 1, panel A and descriptive statistics are reported in Table 1. For manual audiometry, group mean thresholds were in the normal hearing range across all test frequencies (0.25 to 16 kHz) with substantial inter-subject variability in thresholds present at 6 kHz and above. In the lower frequencies (0.25 to 4 kHz) the standard deviation ranged from 4.2 to 5.3 dB. However, the standard deviation increased with test frequency from 9.6 dB at 6 kHz to 18.2 dB at 16 kHz. At most frequencies, group mean thresholds contained 28 ears, however the audiometric threshold at 6 kHz was not measured for three participants (n = 25 ears). Eighteen participants had normal hearing (≤ 15 dB HL) in one ear across all test frequencies from 0.25 to 16 kHz and ten participants had hearing loss that ranged from slight to moderate-severe (slight = 5 ears; mild = 3 ears; moderate = 1 ear; moderate-severe = 1 ear). In the standard frequency region (0.25 to 8 kHz), the degree of hearing loss ranged from slight to mild and in the EHFs it ranged from slight to moderately-severe. Of the ten ears with hearing loss, eight had a sensorineural loss, one was conductive and one was mixed.

**Figure 1.**
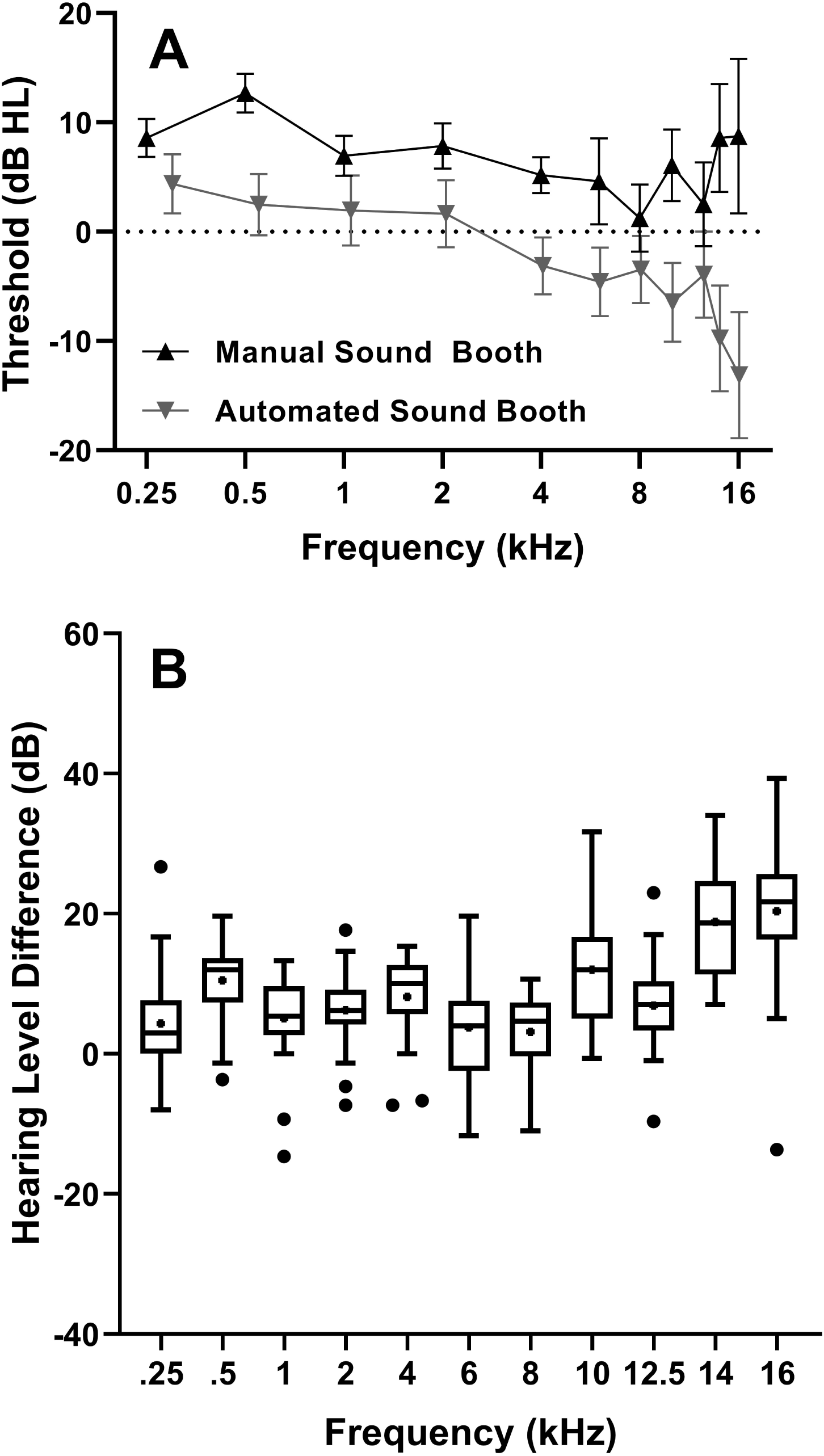
Group mean manual and automated sound booth audiometric thresholds and 95% confidence intervals measured in hearing level (dB HL) for typically developing participants are shown in Panel A. Signed threshold differences between manual and automated sound booth thresholds are in Panel B. Positive values indicate better thresholds on automated audiometry compared to manual audiometry. Conversely, negative values indicate better thresholds on the manual audiometry compared to automated audiometry. Box plots show median (line), interquartile ranges (boxes), 1.5 x interquartile range (whiskers), and outliers (dots).

**Table 1.** Descriptive statistics for audiometric thresholds and reference equivalent threshold sound pressure levels for automated audiometry

|  | **0.25**<br>**kHz** | **0.5**<br>**kHz** | **1**<br>**kHz** | **2**<br>**kHz** | **4**<br>**kHz** | **6**<br>**kHz** | **8**<br>**kHz** | **10**<br>**kHz** | **12**<br>**kHz** | **14**<br>**kHz** | **16**<br>**kHz** |
| --- | --- | --- | --- | --- | --- | --- | --- | --- | --- | --- | --- |
| **Manual Audiometry - Sound Booth** |  |  |  |  |  |  |  |  |  |  |  |
| # of ears | 28 | 28 | 28 | 28 | 28 | 25 | 28 | 28 | 28 | 28 | 28 |
| Mean (SD) | 8.6<br>(4.5) | 12.7<br>(4.6) | 7.0<br>(4.8) | 7.9<br>(5.3) | 5.2<br>(4.2) | 4.6<br>(9.6) | 1.3<br>(7.9) | 6.1<br>(8.4) | 2.5<br>(10.0) | 8.6<br>(12.8) | 8.8<br>(18.2) |
| Median | 10.0 | 15.0 | 5.0 | 10.0 | 5.0 | 5.0 | 0.0 | 5.0 | 0.0 | 5.0 | 5.0 |
| **Automated Audiometry - Sound Booth** |  |  |  |  |  |  |  |  |  |  |  |
| # of ears | 27 | 27 | 27 | 28 | 27 | 26 | 27 | 27 | 27 | 27 | 27 |
| Mean (SD) | 4.4<br>(6.9) | 2.5<br>(7.1) | 2.0<br>(8.2) | 1.6<br>(7.9) | -3.1<br>(6.6) | 0.4<br>(7.8) | -1.5<br>(7.7) | -6.4<br>(9.0) | -3.9<br>(10.0) | -9.8<br>(12.3) | -13.1<br>(14.6) |
| Median | 6.3 | 0.0 | 0.0 | 0.3 | -4.0 | -0.5 | -2.7 | -8.7 | -4.0 | -14.3 | -12.3 |
| **Automated Audiometry – Office** |  |  |  |  |  |  |  |  |  |  |  |
| # of ears | 27 | 28 | 28 | 28 | 27 | 28 | 28 | 28 | 27 | 26 | 26 |
| Mean (SD) | 10.0<br>(6.9) | 5.7<br>(9.2) | 2.5<br>(6.8) | 3.2<br>(7.2) | -1.3<br>(7.4) | 1.9<br>(8.4) | -1.0<br>(8.6) | -6.7<br>(9.4) | -3.8<br>(9.2) | -11.4<br>(11.8) | -11.5<br>(17.0) |
| Median | 8.3 | 6.0 | 2.2 | 3.2 | -1.3 | 0.3 | -2.8 | -6.3 | -7.0 | -13.8 | -14.2 |
| **Automated Audiometry** |  |  |  |  |  |  |  |  |  |  |  |
| RETSPL Values | 15 | 10 | 5 | 5 | 5 | 15 | 18 | 20 | 25 | 35 | 55 |

*Note:* RETSPL = Reference Equivalent Threshold Sound Pressure Level.

For automated audiometry in the sound booth, mean audiometric thresholds were all in the normal hearing range as well. All eighteen participants that showed normal hearing with manual audiometry also had normal hearing with automated sound booth audiometry. However, of the ten ears that had hearing loss with manual audiometry, only five had hearing loss based on automated sound booth thresholds (slight = 4 ears, mild = 1 ear). Similar to manual audiometry, the inter-subject variability from 0.25 to 4 kHz was smaller (SD = 6.6 to 8.2 dB) than in higher frequencies (6-16 kHz; SD = 7.7 to 14.6 dB). For automated sound booth audiometry, there were eleven thresholds from five participants that were not able to be measured because they did not reach threshold convergence in the automated program.

To further evaluate differences in manual and automated sound booth thresholds, signed differences in audiometric thresholds (manual minus automated) are shown in Figure 1, panel B. Positive values indicate better thresholds with automated compared to manual audiometry.

Boxplots show median signed difference values were all greater than 0 dB and ranged from +3 dB at 0.25 kHz to +22 dB at 16 kHz with greater variability at 6 kHz and above, indicating better thresholds with automated audiometry that increased with test frequency.

Absolute differences between manual and automated sound booth thresholds were calculated and used to determine the cumulative counts and percentage of thresholds that were within ±5 dB, ±10 dB, ±15 dB, and > 15 dB from 0.25 to 16 kHz (see Table 2). The percentage of thresholds that were within ±5 dB was extremely low and varied by frequency (mean = 27%, range = 0 to 54%). The percentage of thresholds that were within ±10 dB was only slightly better (mean = 56%, range = 7 to 86%). In general, greater differences between methods were seen within the EHF region, especially at 14 and 16 kHz where less than 30% of thresholds were within ±15 dB.

**Table 2.** Cumulative counts and percentages of the threshold difference between manual and automated audiometry in the sound booth

|  | **0.25**<br>**kHz** | **0.5**<br>**kHz** | **1**<br>**kHz** | **2**<br>**kHz** | **4**<br>**kHz** | **6**<br>**kHz** | **8**<br>**kHz** | **10**<br>**kHz** | **12**<br>**kHz** | **14**<br>**kHz** | **16**<br>**kHz** | **Total** |
| --- | --- | --- | --- | --- | --- | --- | --- | --- | --- | --- | --- | --- |
| **±5 dB** | 15 (54%) | 4 (14%) | 10 (36%) | 9 (32%) | 3 (11%) | 11 (39%) | 14 (50%) | 7 (25%) | 10 (36%) | 0 (0%) | 1 (4%) | 84 (27%) |
| **±10 dB** | 24 (86%) | 10 (36%) | 21 (75%) | 24 (86%) | 14 (50%) | 19 (68%) | 24 (86%) | 10 (36%) | 20 (71%) | 4 (14%) | 2 (7%) | 172 (56%) |
| **±15 dB** | 25 (89%) | 23 (82%) | 27 (96%) | 27 (96%) | 26 (93%) | 21 (75%) | 27 (96%) | 19 (68%) | 25 (89%) | 8 (29%) | 6 (21%) | 234 (76%) |
| **> 15 dB** | 27 (96%) | 27 (96%) | 27 (96%) | 28 (100%) | 27 (96%) | 24 (86%) | 27 (96%) | 27 (96%) | 27 (96%) | 27 (96%) | 27 (96%) | 295 (96%) |
| **Missing** | 1 (4%) | 1 (4%) | 1 (4%) | 0 (0%) | 1 (4%) | 4 (14%) | 1 (4%) | 1 (4%) | 1 (4%) | 1 (4%) | 1 (4%) | 13 (4%) |

*Note:* Displayed values show the number of ears (percentage).

Linear mixed model results showed a significant effect of test method, frequency, and method by frequency interaction. Specifically, automated audiometric thresholds obtained in the sound booth were significantly lower than manual thresholds in the sound booth (F 1(1,27) = 194.6, *p* < 0.001). Audiometric thresholds were elevated in the low frequencies and improved with increasing test frequency (F 1(1,269) = 6.1, *p* < 0.001). The difference in thresholds varied as a function of test frequency and method (F 1(1,269) = 18.6, *p* < 0.001). The least square mean comparisons at each frequency from the linear mixed model are shown in Table 3. In the standard frequencies, automated audiometric thresholds in the sound booth were significantly lower than manual audiometry thresholds at 0.5, 2 and 4 kHz, with mean differences ranging from 6.2 to 10.3 dB. In the EHFs, thresholds at all frequencies (10, 12.5, 14, and 16 kHz) were significantly better for automated compared to manual audiometry with mean differences ranging from 6.6 to 20.7 dB.

**Table 3.**
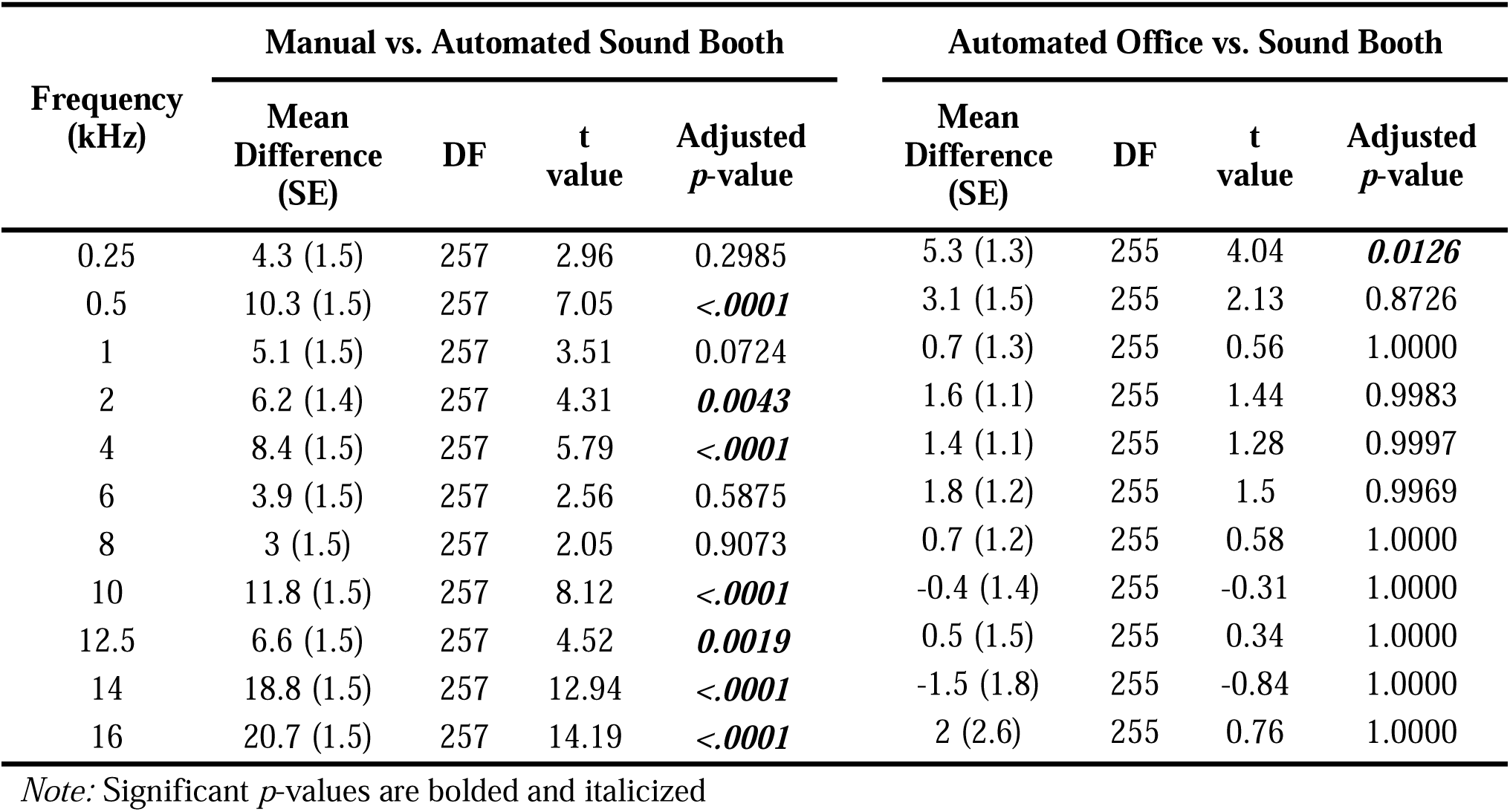
Least square mean comparisons at each test frequency from the linear mixed model

| Frequency<br>(kHz) | Manual vs. Automated Sound Booth |  |  |  | Automated Office vs. Sound Booth |  |  |  |
| --- | --- | --- | --- | --- | --- | --- | --- | --- |
|  | Mean<br>Difference<br>(SE) | DF | t<br>value | Adjusted<br>*p*-value | Mean<br>Difference<br>(SE) | DF | t<br>value | Adjusted<br>*p*-value |
| 0.25 | 4.3 (1.5) | 257 | 2.96 | 0.2985 | 5.3 (1.3) | 255 | 4.04 | ***0.0126*** |
| 0.5 | 10.3 (1.5) | 257 | 7.05 | ***<.0001*** | 3.1 (1.5) | 255 | 2.13 | 0.8726 |
| 1 | 5.1 (1.5) | 257 | 3.51 | 0.0724 | 0.7 (1.3) | 255 | 0.56 | 1.0000 |
| 2 | 6.2 (1.4) | 257 | 4.31 | ***0.0043*** | 1.6 (1.1) | 255 | 1.44 | 0.9983 |
| 4 | 8.4 (1.5) | 257 | 5.79 | ***<.0001*** | 1.4 (1.1) | 255 | 1.28 | 0.9997 |
| 6 | 3.9 (1.5) | 257 | 2.56 | 0.5875 | 1.8 (1.2) | 255 | 1.5 | 0.9969 |
| 8 | 3 (1.5) | 257 | 2.05 | 0.9073 | 0.7 (1.2) | 255 | 0.58 | 1.0000 |
| 10 | 11.8 (1.5) | 257 | 8.12 | ***<.0001*** | -0.4 (1.4) | 255 | -0.31 | 1.0000 |
| 12.5 | 6.6 (1.5) | 257 | 4.52 | ***0.0019*** | 0.5 (1.5) | 255 | 0.34 | 1.0000 |
| 14 | 18.8 (1.5) | 257 | 12.94 | ***<.0001*** | -1.5 (1.8) | 255 | -0.84 | 1.0000 |
| 16 | 20.7 (1.5) | 257 | 14.19 | ***<.0001*** | 2 (2.6) | 255 | 0.76 | 1.0000 |

*Note:* Significant *p*-values are bolded and italicized

Intra-class correlation coefficients were used to examine the reliability or relationship between manual and automated thresholds obtained within the sound booth at each audiometric test frequency (see Table 4). ICC values range between 0 and 1, with higher ICC values indicating a stronger threshold agreement and correlation between the two methods. Results showed poor correlation at most test frequencies with only a moderate correlation at 6, 8, and 12.5 kHz.

**Table 4.** Intra-class correlation coefficients and 95% confidence intervals.

| **Frequency**<br>**(kHz)** | **Manual vs. Automated Sound Booth**<br>**ICC (95% CI)** | **Automated Office vs. Sound Booth**<br>**ICC (95% CI)** |
| --- | --- | --- |
| 0.25 | 0.21 (-0.106, 0.509) | 0.24 (-0.089, 0.540) |
| 0.5 | 0.22 (-0.089, 0.564) | 0.44 (0.096, 0.693) |
| 1 | 0.46 (0.015, 0.738) | 0.52 (0.180, 0.750) |
| 2 | 0.49 (-0.072, 0.785) | 0.82 (0.644, 0.914) |
| 4 | 0.22 (-0.102, 0.553) | 0.69 (0.430, 0.849) |
| 6 | 0.54 (0.189, 0.770) | 0.86 (0.720, 0.936) |
| 8 | 0.71 (0.379, 0.865) | 0.85 (0.707, 0.930) |
| 10 | 0.3 (-0.105, 0.652) | 0.73 (0.494, 0.869) |
| 12.5 | 0.66 (0.002, 0.875) | 0.78 (0.569, 0.893) |
| 14 | 0.38 (-0.067, 0.753) | 0.83 (0.651, 0.918) |
| 16 | 0.42 (-0.090, 0.774) | 0.81 (0.621, 0.910) |

*Note:* <0.5 = poor reliability, 0.5 to 0.75 = moderate reliability, 0.75 to 0.9 = good reliability, >0.9 = excellent reliability

### Automated Audiometry in the Sound Booth vs. the Office

Automated audiometric thresholds measured in both the sound booth and office environment are displayed in Figure 2, Panel A and descriptive statistics are reported in Table 1. For automated audiometry in the office, mean audiometric thresholds were all in the normal hearing range. Mean automated office thresholds were poorest in the low to mid frequencies (3.2 to 10.0 dB) and systematically improved with test frequency from 4 to 16 kHz (–1.3 to –11.5 dB). Similar to manual and automated audiometry in the sound booth, inter-subject variability was present to a lesser degree from 0.25 to 4 kHz (SD = 6.9 to 9.2 dB) and increased with test frequency from 8.4 dB at 6 kHz to 17.0 dB at 16 kHz. There were seven thresholds from four participants that were not able to be measured using the tablet (did not reach convergence). Therefore, the number of ears per frequency varied between 26 and 28 ears.

**Figure 2.**
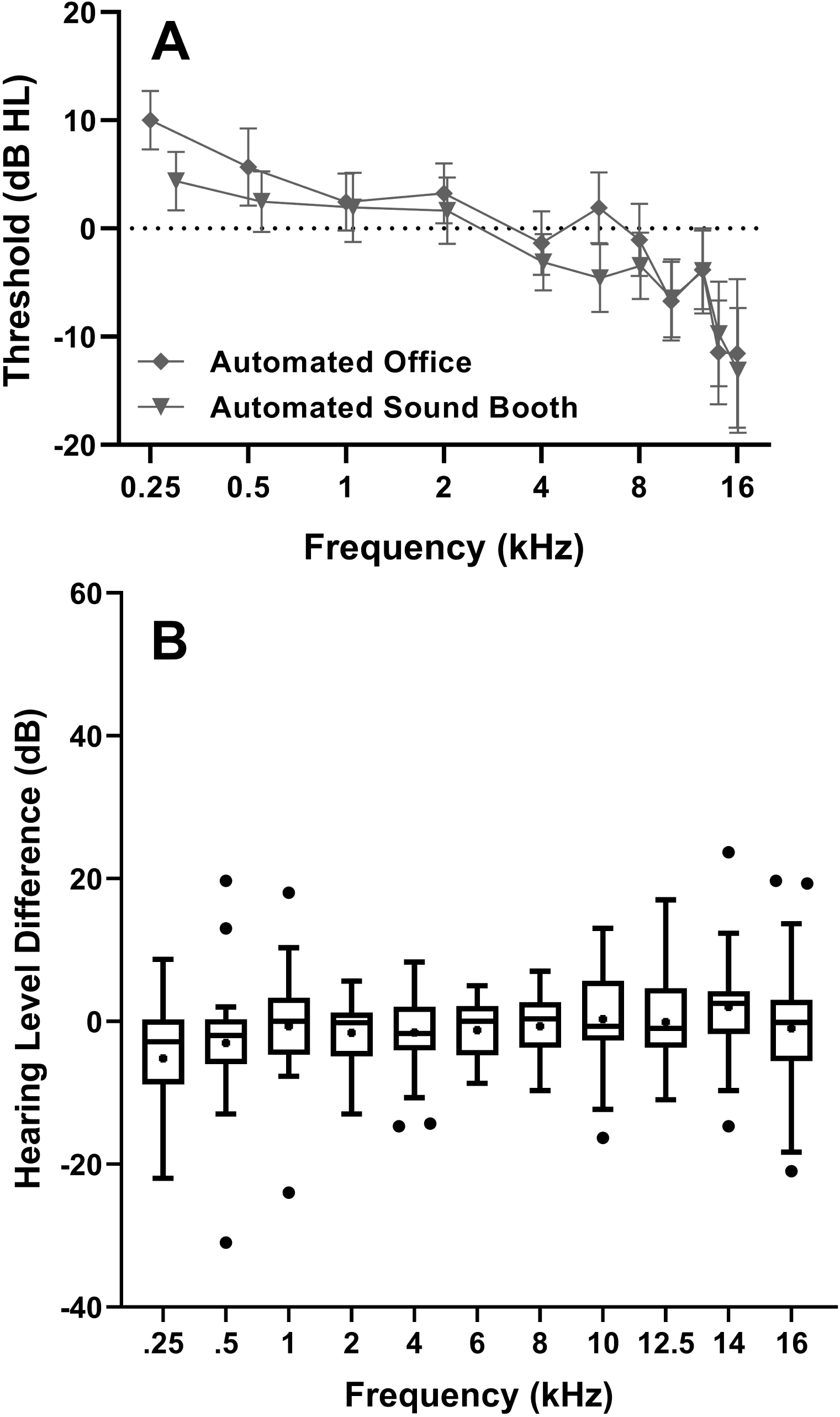
Group mean automated sound booth and office audiometric thresholds and 95% confidence intervals measured in hearing level (dB HL) for typically developing participants are shown in Panel A. Signed threshold differences between automated sound booth and office thresholds are in Panel B. Positive values indicate better thresholds on automated office audiometry compared to automated sound booth audiometry. Conversely, negative values indicate better thresholds on the automated sound booth audiometry compared to automated office audiometry. Box plots show median (line), interquartile ranges (boxes), 1.5 x interquartile range (whiskers), and outliers (dots).

At 0.25 kHz group mean thresholds were 5.6 dB better for the sound booth compared to the office. From 0.5 to 16 kHz, the mean difference in thresholds was minimal and ranged from 0.1 to 3.2 dB. Degree of hearing loss as determined with automated sound booth vs. office thresholds showed twenty participants whose hearing loss category did not change (normal hearing = 18 ears, slight hearing loss = 2 ears). However, there were eight ears whose hearing loss category changed, including 3 ears that improved (slight to normal = 2 ears, mild to slight = 1 ear) and 5 ears that performed worse (normal to slight = 4 ears, normal to mild = 1 ear). To evaluate differences in automated thresholds, signed differences in audiometric thresholds (automated sound booth minus office) are shown in Figure 2, panel B. Positive values indicate better thresholds for the office compared to the sound booth. Conversely, negative values indicate better thresholds in the sound booth compared to the office. Boxplots show median signed difference values that were all around 0 dB (i.e., no threshold difference) and ranged from –2.8 dB to 2.5 dB.

Next, the absolute difference between automated sound booth and office thresholds were calculated and used to determine the cumulative counts and percentage of thresholds that were within ±5 dB, ±10 dB, ±15 dB, and > 15 dB from 0.25 to 16 kHz (see Table 5). Approximately 62% of all thresholds from 0.25 to 16 kHz were within ±5 dB. However, this varied by frequency and ranged from 50% to 71%. A greater percentage of thresholds were within ±10 dB (84%) but ranged from 71 to 96%. In general, the greatest differences between methods was seen at the very lowest (0.25 kHz) and highest (16 kHz) test frequency where less than 80% of thresholds were within ±15 dB.

**Table 5.**
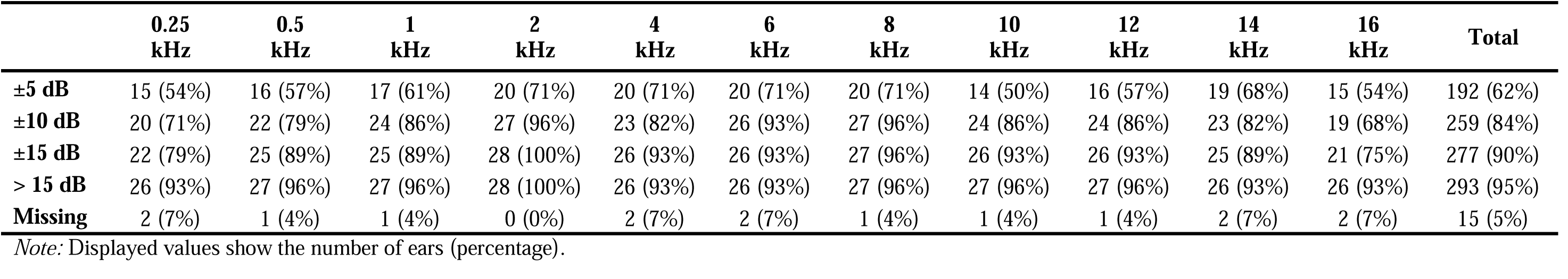
Cumulative counts and percentages of the threshold difference between automated audiometry in the sound booth and office environment.

|  | 0.25<br>kHz | 0.5<br>kHz | 1<br>kHz | 2<br>kHz | 4<br>kHz | 6<br>kHz | 8<br>kHz | 10<br>kHz | 12<br>kHz | 14<br>kHz | 16<br>kHz | Total |
| --- | --- | --- | --- | --- | --- | --- | --- | --- | --- | --- | --- | --- |
| **±5 dB** | 15 (54%) | 16 (57%) | 17 (61%) | 20 (71%) | 20 (71%) | 20 (71%) | 20 (71%) | 14 (50%) | 16 (57%) | 19 (68%) | 15 (54%) | 192 (62%) |
| **±10 dB** | 20 (71%) | 22 (79%) | 24 (86%) | 27 (96%) | 23 (82%) | 26 (93%) | 27 (96%) | 24 (86%) | 24 (86%) | 23 (82%) | 19 (68%) | 259 (84%) |
| **±15 dB** | 22 (79%) | 25 (89%) | 25 (89%) | 28 (100%) | 26 (93%) | 26 (93%) | 27 (96%) | 26 (93%) | 26 (93%) | 25 (89%) | 21 (75%) | 277 (90%) |
| **> 15 dB** | 26 (93%) | 27 (96%) | 27 (96%) | 28 (100%) | 26 (93%) | 26 (93%) | 27 (96%) | 27 (96%) | 27 (96%) | 26 (93%) | 26 (93%) | 293 (95%) |
| **Missing** | 2 (7%) | 1 (4%) | 1 (4%) | 0 (0%) | 2 (7%) | 2 (7%) | 1 (4%) | 1 (4%) | 1 (4%) | 2 (7%) | 2 (7%) | 15 (5%) |

*Note:* Displayed values show the number of ears (percentage).

Linear mixed model results for automated sound booth compared to office did not show a significant effect of test method (F 1(1, 27) = 1.9, *p* < 0.179) but there was a significant effect of test frequency, and method by frequency interaction. Specifically, audiometric thresholds were elevated in the low frequencies and improved with increasing test frequency (F 1(1, 267) = 6.1, *p* < 0.001). The difference in thresholds varied as a function of test frequency and method (F 1(1, 255) = 6.3, *p* < 0.001). The least square mean comparisons at each frequency from the linear mixed model are shown in Table 3. Automated audiometric thresholds obtained in the sound booth were significant better at 0.25 kHz with no significant differences observed from 0.5 to 16 kHz. Across all test frequencies the linear mixed model mean differences were all ≤ 5.3 dB.

Intra-class correlation coefficients and 95% confidence intervals for the automated audiometry sound booth vs. office analysis are shown in Table 5. Results showed poor reliability between methods at 0.25 and 0.5 kHz, with moderate reliability at all other test frequencies from 1 to 16 kHz.

### Automated Office vs. Sound Level Measurements Ambient Noise Measurements

Ambient noise measurements were completed once in each of the sound booths used for manual and automated audiometry. Comparison of these values showed nearly identical values for Leq (Booth A = 31.6 dBA, Booth B = 34.1 dBA), minimum sound level (Booth A = 31.3 dBA, Booth B = 32.2 dBA), and maximum sound level (Booth A = 42.1 dBA, Booth B = 55.5 dBA). In contrast, ambient noise measurements were recorded during office tablet testing for each participant. As anticipated, the sound level measurements from the office showed a higher Leq (median = 55.1 dB A, range = 53.9 to 63.0), minimum sound level (median = 53.3 dB A, range = 49.7 to 54.6) and maximum sound level (median = 69.8 dB A, range = 57.3 to 85.0).

To evaluate the relationship between ambient noise level and automated thresholds measured in the office, Leq (Figure 3.) and maximum sound level (Figure 4) were plotted against automated thresholds at each test frequency. Within each figure, results of the Spearman rank correlation analysis are shown along with linear regression lines. Spearman rank correlation analysis did not show any significant relationships between the Leq and automated thresholds from 0.25 to 16 kHz measured in the office (*p* ≥ 0.118). Similarly, there were no significant relationships between the maximum sound level and automated office thresholds at any test frequency (*p* ≥ 0.059).

**Figure 3.**
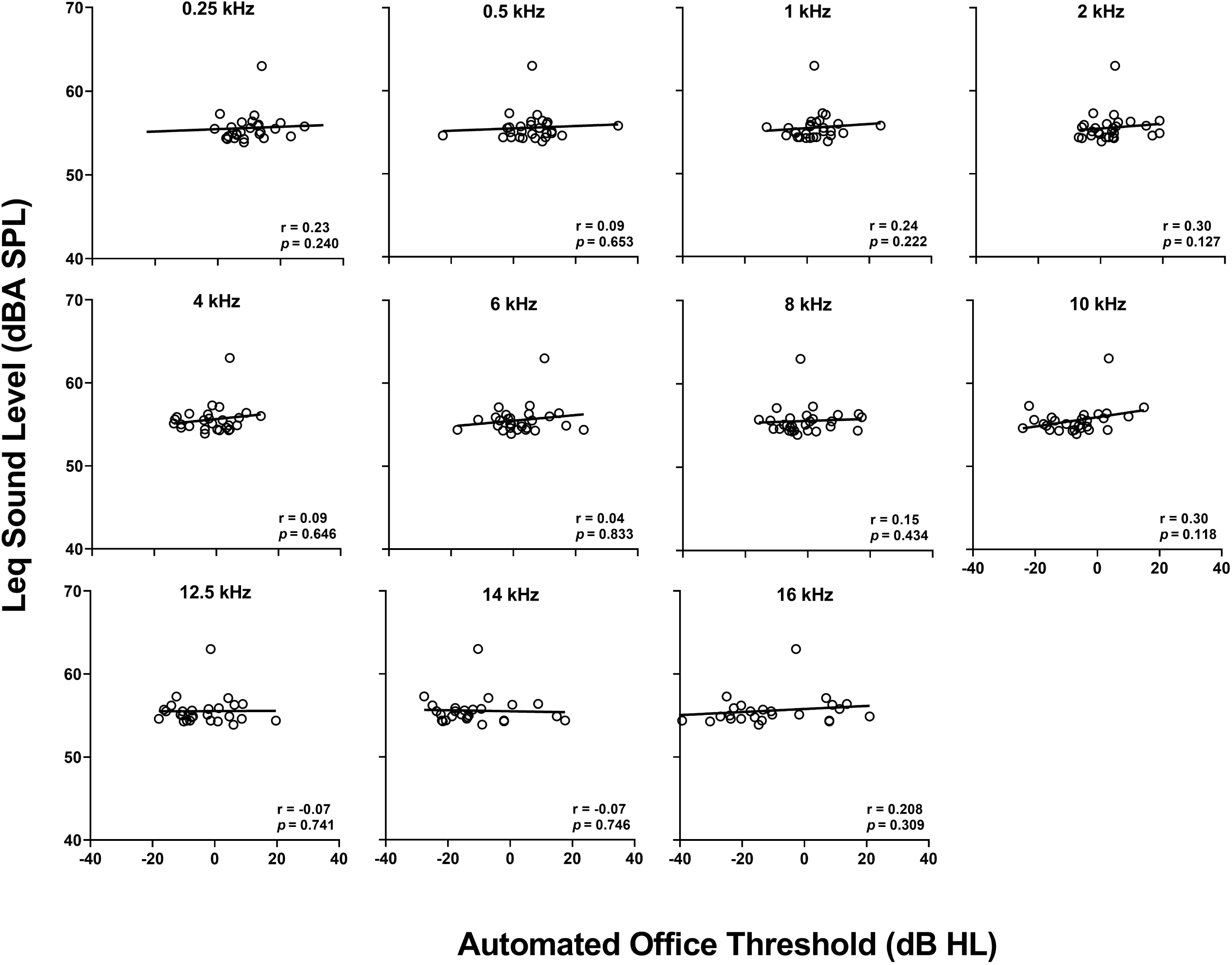
The long-term average (Leq) sound level (dBA) was plotted as a function of automated thresholds measured in the office environment. Regression lines, Spearman rank correlation coefficients, and significance values are displayed in each figure.

**Figure 4.**
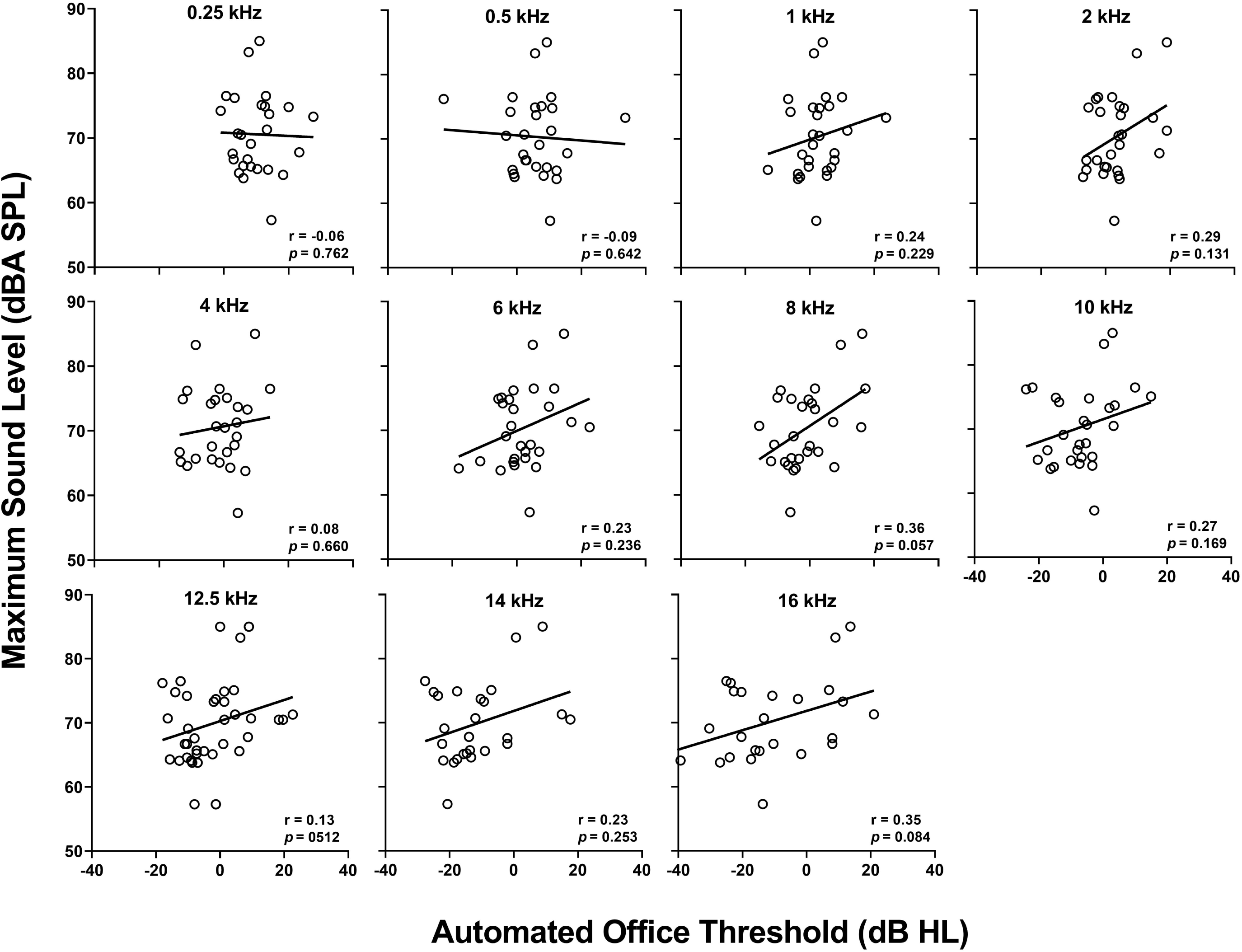
The maximum sound level (dBA) was plotted as a function of automated thresholds measured in the office environment. Regression lines, Spearman rank correlation coefficients, and significance values are displayed in each figure.

## DISCUSSION

There is a substantial body of research showing that automated audiometry is reliable and effective. A meta-analysis on the validity of automated threshold audiometry showed equivalent test re-test reliability between automated and manual audiometry (<u>Mahomed et al., 2013</u>). However of the 29 studies published prior to 2012 that were included in the analysis, only five studies included children, none of the studies evaluated EHF thresholds, and the effect of test location (sound booth vs. non-sound treated room) was not assessed. More recent studies have focused on these important factors to allow hearing health care services to become more accessible to rural and low socioeconomic communities, ototoxicity monitoring at the patient bedside for individuals who are not well enough to travel to the sound booth, and as a screening tool for children in the school setting.

<u>Meinke et al. (2017)</u> assessed test-retest variability of thresholds (.5 to 8 kHz) using the same WAHTS with passive, noise attenuating headphones employed in the present study for 20 workers in six different conference rooms at a workplace, compared to a mobile sound booth. Average automated thresholds obtained with the WAHTS were equivalent to mobile sound booth audiometry at 1, 2, 3 and 8 kHz and within ±5 dB at 0.5, 4, and 6 kHz. In 40 adult participants, <u>Bastianelli et al. (2019)</u> reported that 96% of automated thresholds measured in a quiet exam room using the Shoebox tablet-based audiometer were within ± 10 dB to standard manual thresholds in a sound booth from .05 to 4 kHz. Similarly, <u>Serpanos et al. (2022)</u> showed automated thresholds measured in a clinical exam room using KUDOwave 5000 audiometry were within ± 10 dB for 95% of threshold measurements and within ± 5 dB for ≥ 89% of threshold measurements in 69 adults with normal hearing and hearing loss.

<u>Govender and Mars (2017)</u> studied the KUDUwave 5000 PC-based automated hearing test (0.25 to 8 kHz) in 50 children aged 6 to 13 years in a school and reported that automated air conduction results across the frequency range corresponded with conventional air conduction results within ±5 dB in 81% of ears, and within ±10 dB in 14% of ears, while 5% of ears had a difference greater than ±10 dB. Another study using the KUDUwave system outside a sound booth in adults showed air-conduction thresholds within ± 5 dB in >90% of cases (<u>Swanepoel de et al., 2015</u>). In a study of both children (n = 5) and adults (n = 44), <u>Thompson et al. (2015)</u> compared manual and automated thresholds at .5, 1, 2, and 4 kHz using the Shoebox tablet-based audiometer. Results showed automated thresholds were within 5 dB HL of manual thresholds for 140 of 172 (81%) and within 164 or 172 tests (95%). <u>Magro et al. (2020)</u> used the WAHTS tablet-based audiometry to screen 120 children at 1, 2, and 4 kHz. They reported sensitivity of 100% and specificity 76% for the WHATS tablet in children compared to standard audiometry.

Studies that include EHF audiometry, especially in children, are limited. Shoebox automated audiometry was recently studied to screen for EHF hearing loss in adults with cystic fibrosis at risk for ototoxicity (<u>Vijayasingam et al., 2020</u>). That study reported that automated audiometry in adults was equivalent to standard audiometry, with 93% sensitivity and 88% specificity. Overall, the consensus in previous studies is that tablet audiometry can be used as a reliable tool but needs to be verified with standard audiometry.

The current study is the first concerning EHF threshold automated audiometry in children and adolescents. Strengths are the repeated measure design in both a sound booth and office environment, as well as comparisons with manual threshold audiometry in a sample that included a range of hearing levels. Overall, eighteen participants that showed normal hearing with manual audiometry also had normal hearing with automated sound booth audiometry (100% specificity). However, of the ten ears that had hearing loss with manual audiometry, only five ears had hearing loss based on automated sound booth thresholds (slight = 4 ears, mild = 1 ear), or 50% sensitivity. The strict criterion for hearing loss (>15 dB at any frequency) means that a small adjustment in criterion would affect test sensitivity. Automated threshold reliability (sound booth versus office) was within ±5 dB 62% of the time and was cumulatively within ±10 dB 84% of the time.

When examining the data, substantial variability in extended high frequency thresholds can be seen across participants in Figures 1 and 2. The variability may be related to the population studied, high inter-subject variability observed in EHF threshold even in individuals with normal hearing (<u>Rodríguez Valiente et al., 2014</u>), or the presence of standing waves within the ear canal that may affect inter-subject variability in the EHF region. For example, in children and adults, <u>Schmuziger et al. (2004)</u> reported decreased test-retest reliability at 14 and 16 kHz (83-87% of thresholds within ±5 dB) compared to 0.5-12.5 kHz (90-99% of thresholds within ±5 dB). Furthermore, <u>Beahan et al. (2012)</u> reported a significant effect of age on the test-retest reliability of EHF thresholds. Children between 10-13 years of age showed the best test-retest reliability with ≥ 96% of thresholds within ±10 dB which decreased to 86-94% for children between 4-6 years of age.

Overall, our results suggest that WAHTS automated audiometry in children can be done outside of a sound booth with good reliability, thereby making ototoxicity monitoring more feasible on a routine basis. Additionally, our results show good reliability between automated thresholds obtained in the booth vs. office environment from 6-16 kHz (ICCs from .73 to .86), which is the frequency range most important for ototoxicity monitoring. Despite the lack of a significant relationship between ambient noise measurements and automated thresholds, our results show poor reliability between automated sound booth and office thresholds at 0.25 and 0.5 kHz, frequencies that are most affected by ambient noise. Lastly, results from the current study would only apply to the WAHTS because the sound isolation algorithms within the circumaural transducers is a pretty important determinant of the automated thresholds obtained in the office environment.

### Limitations and Future Directions

First, limiting environmental noise interference may be addressed by a built-in sound meter that pauses testing until the sound level is within the acceptable range (<u>Vijayasingam et al., 2020</u>). Second, differences in reliability between the current study and those in adults may be related to poorer attention in children. In general, children and teens aged 10-18 years were able to complete automated audiometry within 15 minutes, despite occasional environmental noise and distractions. Most children did not have any issues while taking the test, but some younger participants struggled with attention and needed redirection from the examiner. Shortening the testing protocol to five frequencies as suggested by <u>Fausti et al. (1992)</u> as well as inclusion of photos or an interactive children’s character with positive reinforcement for a correct response could help reduce the child’s fatigue to the task. Third, EHF audiometry shows potential for greater sensitivity to detecting hearing loss, but intra-subject variability is higher due to the greater presence of standing waves in the patient’s ear canal (<u>Lee et al., 2012</u>). Lastly, for automated sound booth audiometry, there were eleven thresholds from five participants that were not able to be measured (did not reach convergence). This is an important reliability feature of automated audiometry that is not quantified in standard audiometry, thus false positive and negative responses, and lapses in attention can be more easily detected using automated audiometry.

Limitations with the testing equipment included that the children sometimes reported headphones to be heavy or uncomfortable, which was lessened by giving the child breaks between ears. Development of lighter, pediatric sized headphones would be helpful. An additional concern was that some younger children did not understand the printed directions. Encouraging the children to re-read the instruction page or providing verbal directions solved this problem. Further directions could include developing a recorded version of the directions that could be played directly into the headphones and could be replayed if they need re-instruction. Additionally, in order to establish WAHTS audiometry as a practical clinical tool for ototoxicity monitoring, a large scale pediatric validation study is needed to establish true positive and negative rates in ototoxicity.

In summary, we found that automated audiometric thresholds can be expected to be approximately 5 dB HL better than manual audiometric thresholds based on previous literature (<u>Meinke et al., 2017</u>), and confirmed in children in this study, at least for 0.5 to 8 kHz. We found substantially better thresholds with automated audiometry above 8 kHz. Due to differences in headphones and methodology, age-referenced normative values are needed for automated audiometry specific to the type of circumaural earphones employed.

## CONCLUSIONS

The WAHTS wireless automated tablet-based hearing test system was used to evaluate the EHF hearing levels for typically developing children and adolescents, mostly without reported hearing problems. Results showed large variability and threshold differences between manual and automated audiometry within a sound booth, mainly in the EHFs. However automated audiometry results obtained in a sound booth and in an office and were found to agree within ±10 dB the majority of the time. Automated audiometry could prove to be useful for ototoxicity monitoring, once a larger normative sample has been collected. Improvements in ease of use for children would advance the probability of this equipment becoming readily used by clinicians.

## Data Availability

All data produced in the present study are available upon reasonable request to the authors

## Abbreviations

(WAHTS): Wireless Automated Hearing Test System
(EHF): Extended High Frequency

## Acknowledgements and Author Contributions

Portions of this study were presented at the American Auditory Society 2019 and the American Academy of Audiology 2019. The content of this manuscript is solely the responsibility of the authors and does not reflect the official views of Cincinnati Children’s Hospital Medical Center or Decibel Therapeutics. Appreciation is extended to the participants and their parents who participated in this research.

*C.M.B and L.M.B performed experiments, analyzed data, and cowrote the paper. T.Q. and E.L. designed experiments and provided critical revision to the manuscript. L.L. provided statistical analysis. L. L. H. designed and performed experiments, analyzed data, and cowrote the paper*.

**Supplemental Digital Content.** Shown is the instruction screen of the WAHTS automated audiometry test (left), and corresponding first frequency showing ear being tested and large response button (right).

## Notes

**Financial Disclosures/Conflicts of Interest**: The authors have no conflicts of interest to disclose. This research was supported by Decibel Therapeutics (Hunter) and the National Institute of Health, Clinical and Translational Science Award Program Grant 5UL1TR001425-04 (Center for Clinical and Translational Science Training at the University of Cincinnati)

### Competing Interest Statement

The authors have declared no competing interest.

### Funding Statement

This research was supported by Decibel Therapeutics (Hunter) and the National Institute of Health, Clinical and Translational Science Award Program Grant 5UL1TR001425-04 (Center for Clinical and Translational Science Training at the University of Cincinnati).

### Author Declarations

The Cincinnati Childrens Hospital Medical Center Institutional Review Board gave ethical approval for this work.

