## Supplementary figures and images for "Extended High-Frequency Audiometry using the Wireless Automated Hearing Test System Compared to Manual Audiometry in Children and Adolescents"

### Supplemental Table 1

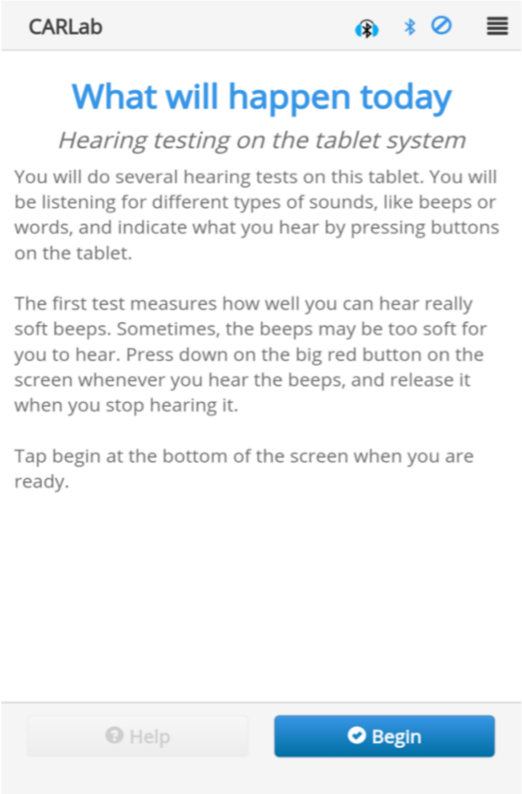

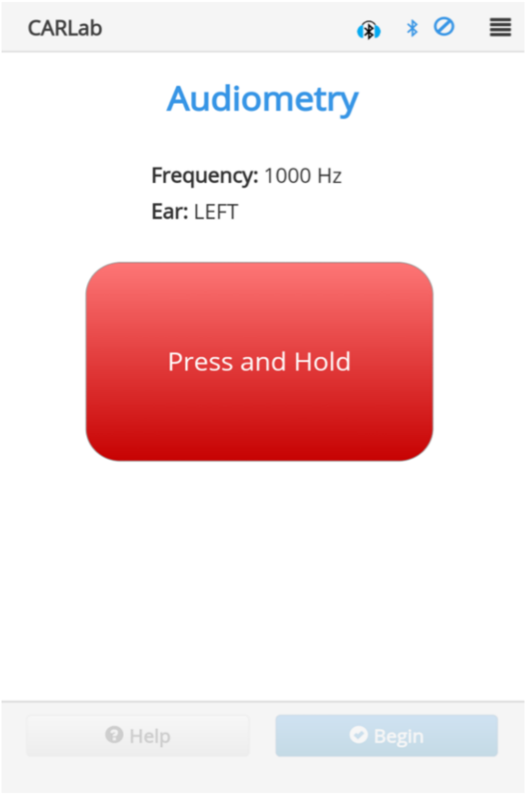
